# Neural synchrony reflects pain and co-occurring psychological symptoms: a transdiagnostic magnetoencephalography study using multivariate modeling

**DOI:** 10.1101/2024.11.15.24317356

**Authors:** Majid Saberi, Matthew Ventresca, Rouzbeh Zamyadi, Mryam Ali, Jing Zhang, Oshin Vartanian, Rakesh Jetly, Venkat Bhat, Shawn G Rhind, J Don Richardson, Benjamin T Dunkley

## Abstract

**Objective:** Pain commonly co-occurs with psychological symptoms, yet the neural synchrony patterns associated with this shared symptom burden remain incompletely understood. We examined whether frequency-specific neural synchrony is associated with pain severity and co-occurring psychological symptoms in a transdiagnostic military cohort with heterogeneous symptom presentations.

**Methods:** Resting-state magnetoencephalography data were acquired from military personnel and veterans exhibiting varying levels of pain, anxiety-, depression-, and PTSD-related symptoms. Frequency-specific neural synchrony was estimated between regions within literature-derived pain-relevant brain networks. Multivariate partial least squares regression was used to model associations between neural synchrony patterns and pain severity, as well as composite measures reflecting joint pain and psychological symptom burden.

**Results:** Pain severity was associated with distributed synchrony patterns, with the strongest associations observed in the beta and high-gamma frequency bands. Joint pain-anxiety symptom burden was primarily associated with theta- and gamma-band synchrony, whereas joint pain-depressive and pain-PTSD symptom burden showed predominant associations within the gamma band, with fewer beta-band effects. Across symptom domains, overlapping but frequency-specific synchrony patterns were identified across distributed brain networks.

**Conclusions:** Pain severity and co-occurring psychological symptoms are associated with partially overlapping, frequency-specific neural synchrony patterns involving distributed brain networks.

**Significance:** These findings support a network-level neurophysiological framework for understanding pain and psychological symptom co-occurrence and highlight frequency-specific MEG synchrony patterns as potential markers of multidimensional symptom burden.

**Highlights:**

– MEG synchrony patterns associate with pain severity across beta and high-gamma bands
– Pain-anxiety symptom burden links to theta- and gamma-band synchrony
– Pain-depression and pain-PTSD burden show predominant gamma-band synchrony

## 1. Introduction

Pain is a multidimensional experience shaped by sensory, emotional, and cognitive processes, and its severity is closely linked to broader psychological functioning [1]. Across populations, higher pain levels often co-occur with elevated anxiety-, depressive-, and post-traumatic-stress symptoms [2–7], reflecting shared vulnerability across affective and pain-processing systems [8–12]. Understanding how neural dynamics relate to this co-occurring symptom burden is essential for clarifying mechanisms that contribute to impaired functioning and treatment challenges.

Chronic pain, defined as pain persisting for over three months [13], involves physiological and psychosocial factors affecting daily life [1]. It impacts approximately 20.5% of adults in the United States [14] and 18.9% in Canada [15], imposing significant socioeconomic burdens [16]. These observations underscore the importance of investigating brain-based mechanisms that contribute to variability in pain severity and its psychological correlates.

Chronic pain is driven by complex neural mechanisms in the peripheral and central nervous system. Peripheral sensitization triggers pain signals via inflammatory mediators, while central sensitization processes in the spinal cord and brain sustain them [17,18]. Glial cells release pro-inflammatory cytokines, amplifying pain signals [19], and dysfunction in descending modulatory pathways further exacerbates pain persistence [7,20]. Together, these mechanisms highlight the relevance of examining central neural dynamics associated with pain severity.

Neuroimaging research demonstrates that pain engages distributed brain regions involved in somatosensory processing, salience detection, emotional regulation, and cognitive control [21–24]. Guided by this literature, rather than adopting a whole-brain exploratory approach, we employed a hypothesis-driven set of pain-relevant brain regions, the “dynamic pain connectome” [25,26], that has been consistently implicated across experimental and clinical pain studies [27–29].

Magnetoencephalography (MEG) provides a unique opportunity to examine frequency-specific neural synchrony, a key marker of large-scale communication across brain networks. Prior work has demonstrated that pain severity is associated with alterations in synchrony across widespread frequency bands, particularly within salience, somatosensory, and default mode networks [27–29,30]. However, little is known about how these frequency-specific synchrony patterns relate to the joint expression of pain and psychological symptoms within individuals.

Psychological symptoms, including anxiety-, depressive-, and post-traumatic-stress symptoms, vary dimensionally across individuals and frequently co-occur with higher pain severity [2,3,8–10,5,7]. A transdiagnostic, dimensional framework therefore allows for a more precise characterization of shared neural mechanisms, without relying on categorical diagnostic distinctions [31,32].

In this study, we examined whether frequency-specific MEG synchrony patterns within pain-relevant brain regions are associated with pain severity and co-occurring psychological symptoms. Using multivariate partial least squares regression (PLSR) [33–35], which is well suited for high-dimensional and collinear neural data, we modeled distributed synchrony patterns associated with pain severity as well as composite measures capturing the joint expression of pain with anxiety-, depression-, and PTSD-related symptoms. This approach enables identification of shared neural dynamics underlying overlapping symptom dimensions.

By focusing on a sample recruited from active military members and Veterans with heterogeneous pain and psychological symptom profiles, this study aims to clarify how multi-band neural synchrony reflects shared symptom burden in a population characterized by substantial clinical variability. This transdiagnostic, dimensional perspective provides insight into neural mechanisms that may contribute to the frequent co-occurrence of pain and psychological symptoms.

## 2. Methods

### 2.1. Participants

We recruited n = 106 active-duty military personnel and Veterans of the Canadian Armed Forces (CAF) through a combination of community outreach, public postings/flyers, and clinical referrals from Operational Stress Injury (OSI) clinics, as part of a cross-sectional observational study examining the relationships between brain, cognitive, and mental health outcomes in military.

This study did not target a specific chronic pain condition or any particular mental health diagnosis; instead, recruitment aimed to capture the heterogeneous pain experience and psychological symptoms levels commonly observed in this population, enabling a dimensional and transdiagnostic characterization of symptom variability.

PTSD was not required for participation; while some participants were recruited through clinical PTSD treatment pathways, diagnostic status for those individuals was confirmed by structured clinical interview, whereas psychological symptoms in the broader sample were characterized using validated self-report scales rather than categorical diagnostic assessments.

After quality assurance procedures, we retained n = 99 usable sets of individual data (mean age = 46.8 years, SD = 9.8 years; 88% male, n = 87). From the 106 initial sets, three were removed due to the inability of the participants to complete either MEG or MRI scans for various reasons, such as scanner malfunction, or participants not being able to fit in the scanner(s). In addition, two sets were removed during preprocessing, due to excessive sensor noise. Another two participants were removed due to incomplete questionnaire data.

The participants served in a wide variety of capacities across numerous roles including infantry, armoured, ordnance disposal, special operations forces, paratroopers, artillery, and administrative or technical roles. For participants with confirmed mental health diagnoses (e.g., PTSD), diagnostic status was established through structured clinical interview conducted by a qualified clinician; however, diagnostic information was available only for a subset of the sample, and therefore all analyses used dimensional symptom scores rather than diagnostic categories.

Inclusion criteria were being between the ages of 20 and 65 and the ability to be imaged in an MRI scanner’s confined space. Exclusions were made for individuals with ferrous metal implants that could affect MEG or MRI scanning, or those with a history of seizures, or active substance use disorders. Substance use was screened via the Alcohol Use Disorders Identification Test (AUDIT) and a brief substance use questionnaire; while these measures were not used for formal diagnosis, participants with active substance use disorder were excluded.

Written informed consent was obtained from all participants, and the Hospital for Sick Children Research Ethics Board approved the study protocol. All participants provided their written informed consent for study participation.

### 2.2. Data Collection

Five minutes of eyes-open resting-state MEG data (600 Hz) in the supine position was collected using a 151-channel CTF system at the Hospital for Sick Children in Toronto. Fiducial coils, placed at the left and right pre-auricular points as well as the nasion, were used for continuous head motion monitoring. For MEG source modelling, MRI scans were collected on a 3T Siemens PrismaFit scanner equipped with a 20-channel head and neck coil. A high-resolution 3D MPRAGE T1-weighted image was obtained with an isotropic voxel size of 0.8 mm (TR of 1870 ms, TE of 3.1 ms, and TI of 945 ms) over a field of view measuring 240 × 256 mm and comprising 240 slices each 0.8 mm thick.

On the day of scanning, participants completed self-reported screening questionnaires for psychiatric, neurological, neurobehavioural, neurocognitive, and pain symptoms. These included the long-form McGill Pain Questionnaire [36], which provided a total Pain Rating Index (PRI; range 0-78) and included checklist items on pain location and qualitative descriptors of pain sensation. In this version, participants select one descriptor from each of 20 categories of pain. Within each category, descriptors are ranked in ascending intensity. The PRI is then calculated as the sum of all selected category scores, yielding a total range of 0-78. Specific clinical pain diagnoses (e.g., neuropathic, musculoskeletal, migraine) were not systematically collected; therefore, pain-related analyses rely on self-reported symptom severity rather than formal diagnostic coding.

Psychological symptoms were assessed using the Generalized Anxiety Disorder (GAD-7) scale [37], a 7-item measure scored from 0 (“not at all”) to 3 (“nearly every day”), yielding a total range of 0-21; the total score was used as a dimensional index of anxiety symptoms. Depression symptoms were assessed using the Patient Health Questionnaire-9 (PHQ-9) [38], a 9-item scale scored 0-3 per item, with a total range of 0-27; the total score served as the dimensional depression variable. PTSD-related symptoms were assessed using the PTSD Checklist-Military Version (PCL-M) [39], which is based on DSM-IV criteria to maintain consistency with prior research, and includes 17 items scored from 1 (“not at all”) to 5 (“extremely”), with a total range of 17-85; the total score was used as the dimensional PTSD symptom variable.

### 2.3. MEG Signal Processing

MEG data were processed using FieldTrip and SPM toolboxes [40,41]. Data were bandpass filtered (1-150 Hz), and epoched into 10-second epochs. On average, 23 trials per participant (range: 13-27, corresponding to 86% of the maximum 27 trials) were retained after artifact rejection. The maximum number of trials were retained after removing trials where head position deviated more than 5 mm from the original position. In addition, epochs including SQUID jumps in excess of ±2 pT were also removed. Subsequently, cardiac and ocular artefacts were removed following independent component analysis (ICA) using the “fastica” method from FieldTrip, and visual inspection of the resulting components.

Source localization was performed to pinpoint cortical activity within thirty-six regions of interest (ROIs) selected a priori based on prior theoretical and empirical work describing a distributed set of brain regions consistently involved in pain perception, modulation, and affective-cognitive processing, often referred to as the “dynamic pain connectome” [25,26]. These ROIs were used as a hypothesis-driven framework for examining frequency-specific synchrony in pain-relevant circuits rather than as a fixed or universal atlas. To localize sources, a forward solution was created for each participant by using the individual anatomical T1-weighted MRI scans. After co-registration of the MEG data to the MRI, a linearly constrained minimum variance (LCMV) beamformer [42] was employed to estimate electrophysiological activity at the predetermined ROIs, using the epoched, artifact-free sensor-level data.

For connectivity analysis, the broadband, source-level time-series data were bandpass filtered into the canonical frequency bands: delta (1-3 Hz), theta (4-7 Hz), alpha (8-14 Hz), beta (15-30 Hz), low-gamma1 (30-55 Hz), low-gamma2 (65-80 Hz), and high-gamma (80-150 Hz). We followed band definitions used in prior MEG studies of pain and neuropsychiatric populations [43,44], to maintain comparability with existing literature from our group and others. The Weighted Phase-Lag Index (wPLI) was used to quantify functional connectivity between pairwise ROI connections [45]. A 36-by-36 connectivity matrix was constructed for each frequency band and epoch per participant by calculating the wPLI between all possible ROI pairs. These matrices were subsequently averaged across epochs to yield a functional connectivity matrix per participant for each frequency band.

For interpretability, the 36 ROIs were grouped into eight canonical large-scale networks (central executive, default mode, salience, somatomotor, subcortical, limbic, ventral attention, and visual). This classification was based on established network atlases and prior literature on the pain connectome, combined with anatomical projection of each ROI. The full list of ROI-to-network assignments is provided in Supplementary Table 1.

### 2.4. Partial Least Squares Modeling

We employed Partial Least Squares Regression (PLSR) [33–34] to model the complex relationships between complete set of neural synchronies within pain-relevant ROIs (630 connections) as independent variables and specific outcomes, including pain severity, and composite variables indexing the joint expression of pain severity with psychological symptoms (pain × anxiety, pain × depression, and pain × PTSD-related symptoms), as dependent variables. Each composite was constructed as the product of standardized pain severity and the corresponding standardized symptom score to capture shared symptom burden, rather than to test formal statistical interactions. Each composite term was modeled separately to avoid conflating variance across anxiety-, depression-, and PTSD-related symptoms, thereby preserving their distinct contributions while still allowing examination of co-occurring symptom dimensions.

PLSR is well-suited to highly collinear data, high-dimensional predictor spaces, and scenarios where the number of predictors exceeds the number of observations. Prior to modeling, neural synchrony values were residualized for age and sex using multiple linear regression to remove potential confounding effects.

Model significance was assessed via permutation testing (10,000 resamples), in which subject labels were shuffled and the entire PLSR re-estimated for each resample. This produced an empirical null distribution against which the observed model coefficients were compared to obtain p-values. This procedure provides a global assessment of whether the multivariate pattern linking synchrony to symptom scores exceeds chance-level covariance [35]. Details on statistical inference and multiple comparison considerations are provided in the Statistical Analysis section.

Because our primary aim was to interpret the model coefficients linking neural synchrony to pain and mental health outcomes, PLSR modeling were conducted separately for each canonical frequency band and outcome variable. This frequency-specific approach allowed us to examine which oscillatory processes contributed to shared symptom variability without assuming common effects across rhythms.

### 2.5. Statistical Analysis

All statistical procedures were pre-specified to address the distributional properties of the data and the high dimensionality of the MEG connectivity features. Symptom measures (pain severity, anxiety-, depression-, PTSD-related symptoms) were inspected for normality, and due to positive skew, non-parametric tests were employed where appropriate. Pairwise associations between clinical measures were assessed using Spearman’s rank correlations, with significance evaluated using two-tailed tests. False discovery rate (FDR) correction was applied to control for multiple comparisons across correlations.

All statistical procedures were pre-specified to address the distributional properties of the data and the high dimensionality of the MEG connectivity features. Symptom measures (pain severity; anxiety-, depression-, and PTSD-related symptoms) were inspected for normality, and due to positive skew, non-parametric tests were employed where appropriate. Pairwise associations between clinical measures were assessed using Spearman’s rank correlations, with significance evaluated using two-tailed tests. False discovery rate (FDR) correction was applied to control for multiple comparisons across correlations.

For modeling the relationship between connectivity and clinical outcomes, PLSR was chosen given its suitability for high-dimensional, collinear predictors relative to sample size. To assess significance of model coefficients, permutation testing (10,000 resamples) was conducted, generating empirical null distributions. Because statistical inference in PLSR is performed on the full multivariate pattern rather than on each variable independently, only a single global test is conducted per model, and additional correction for multiple comparisons is not required [35].

We used the R programming environment and packages “pls” and “caret” for statistical modeling [46–48], along with “ggplot2” and “BrainNet Viewer” for visualization [49,50].

## 3. Results

### 3.1. Pain severity moderately correlates with psychological symptoms

Based on McGill Pain Questionnaire body map responses, the most frequently reported pain locations were the torso/back (65 participants, 66%), neck/shoulders (58 participants, 59%), and lower limbs (58 participants, 59%), followed by the head/face (39 participants, 39%) and upper limbs (25 participants, 25%). Many participants indicated more than one location. These self-reported classifications reflect participant perception of pain origin and do not constitute formal clinical diagnoses.

The distributions of pain severity and anxiety-, depression-, and PTSD-related symptoms were positively skewed, with a substantial number of participants reporting minimal or no symptoms (**Figure 1A**), consistent with the heterogeneous symptom presentation expected in a naturalistic military/Veteran cohort containing both clinical-range and subclinical values. Non-parametric statistics were therefore used for all pairwise associations.

**Figure 1.**
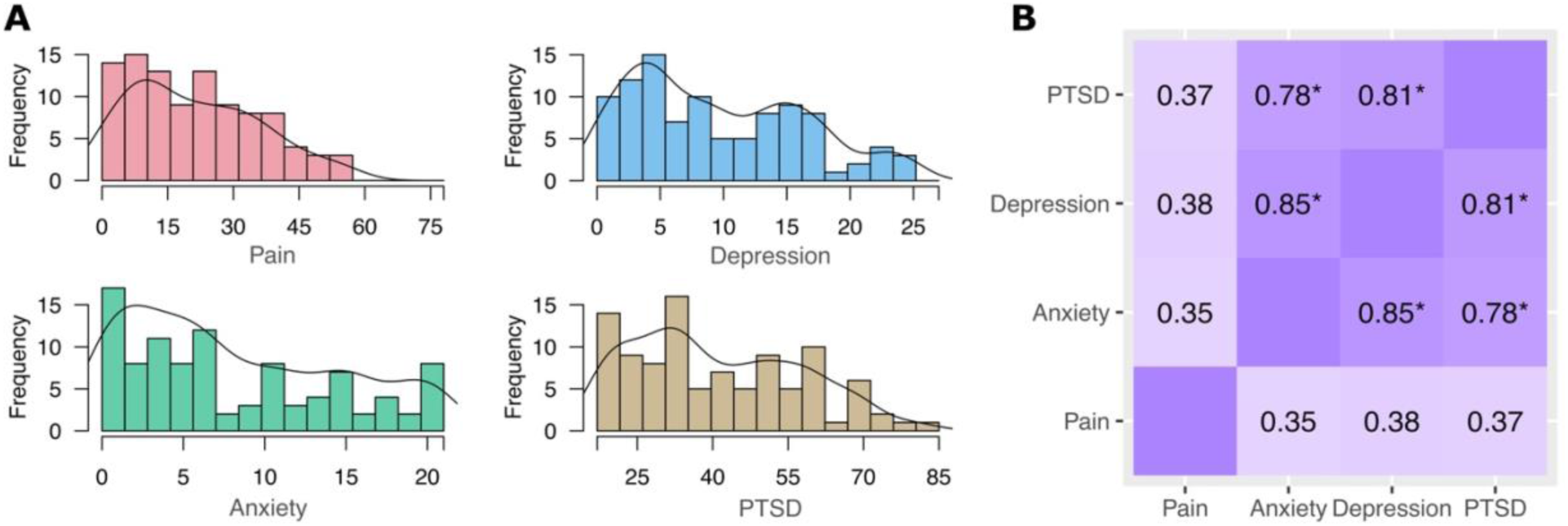
Pain and psychological symptom severity measures and their associations. **(A)** Histogram of pain severity and anxiety-, depression-, and PTSD-related symptom severity across subjects. **(B)** Spearman’s correlation matrix of pain severity and psychological symptom measures. All correlations are statistically significant after multiple comparison correction. Asterisks denote correlation coefficients with large effect sizes; small effect (r = 0.10-0.29); medium effect (r = 0.30-0.49); large effect (r ≥ 0.50).

We employed non-parametric statistics to address these properties. Spearman correlations between pain severity and mental health outcomes were generally low to moderate (all p-values were significant after multiple comparisons with medium effect sizes: 𝑝_pain & anxiety_ = 3.92e-4, 𝑝_pain & depression_ = 1.11e-4, 𝑝_pain & depression_ = 1.31e-4) (**Figure 1B**). In contrast, correlations among psychological symptom measures themselves were high, reflecting substantial shared variance across anxiety-, depression-, and PTSD-related symptoms (all p-values significant after multiple comparisons with large effect sizes).

These moderate associations indicate that pain severity and psychological symptoms are related but not redundant constructs, supporting the use of composite variables to examine their joint expression. Because psychological symptoms share substantial variance, modeling each pain × symptom composite separately allows us to capture distinct dimensions of shared symptom burden without assuming that all psychological domains exert identical neural associations. This dimensional variation motivates the subsequent multivariate analyses examining frequency-specific neural synchrony patterns in relation to pain and co-occurring psychological symptoms.

### 3.2. Frequency-specific neural synchrony in the pain connectome reflects pain severity and co-occurring psychological symptoms

Given the non-parametric distribution of symptom measures, their high intercorrelation, and the collinear nature of the connectivity data, partial least squares regression (PLSR) was used to model the relationships between frequency-specific neural synchrony between pain-relevant brain regions and continuous clinical measures.

#### 3.2.1. Pain severity is associated with frequency-specific neural synchrony

Significant associations between pain severity and neural synchrony were identified across many connections in the pain connectome, particularly in the beta and high-gamma frequency bands (**Figure 2, Row 1**).

**Figure 2.**
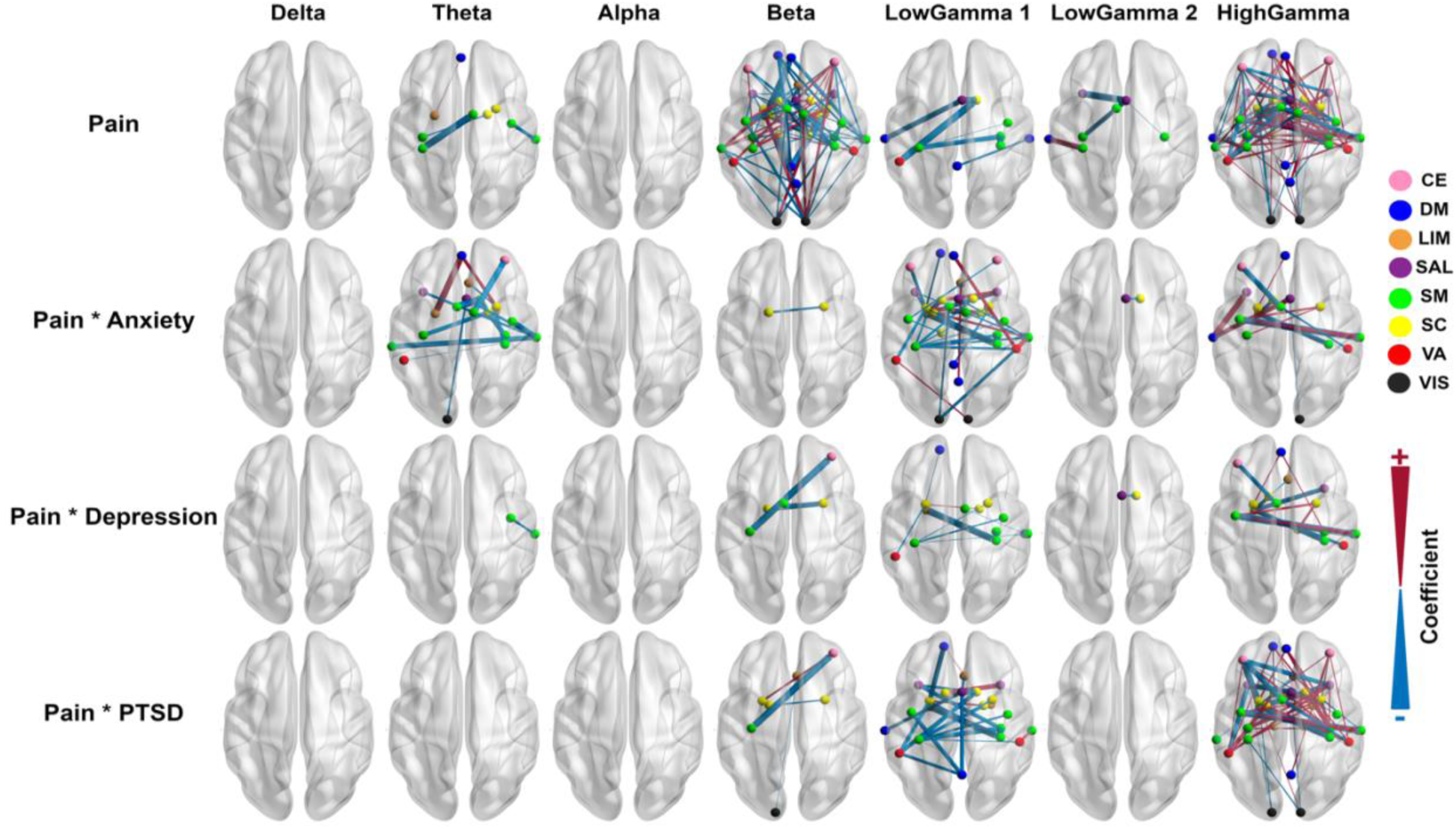
Frequency-specific associations between neural synchrony, pain severity, and co-occurring psychological symptom burden. Brain maps show connections whose regional synchrony was significantly associated with pain severity and with composite measures indexing the joint expression of pain severity with anxiety-, depression-, and PTSD-related symptoms across frequency bands, as identified by PLSR. Rows correspond to pain severity alone and to pain-symptom composite scores, reflecting shared symptom burden rather than formal statistical interactions. Columns correspond to canonical frequency bands (delta to high gamma). Nodes represent regions from a literature-derived set of pain-relevant brain regions and are color-coded by large-scale network membership (see Supplementary Table 1 for full ROI-to-network mapping). The color of the links denotes the direction of the association (red indicates positive associations; blue indicates negative associations), while link width reflects the magnitude of the PLSR coefficient. Abbreviations: CE: Central Executive, DM: Default Mode, LIM: Limbic, SAL: Salience, SM: Somatomotor, SC: Subcortical, VA: Ventral Attention, VIS: Visual.

Theta-band synchrony exhibited an inverse relationship with pain severity across three connections in the somatosensory and subcortical networks, in contrast to the positive association of the left amygdala to the medial prefrontal cortex.

Beta-band synchrony showed large-scale associations with pain severity, exhibiting both positive and negative relationships. Connections involving the default mode and somatosensory networks were predominantly negatively associated with pain severity, whereas connections involving the central executive and subcortical networks showed the greatest number of positive associations with pain severity.

Across the low-gamma frequency ranges, several within-network and between-network connections were negatively associated with pain severity, with the exception of a positive strong association between the left primary somatosensory cortex and the left medial temporal lobe in the low-gamma2 band.

In the high-gamma band, synchrony between regions involving the central executive network, default mode network, and subcortical structures was predominantly positively associated with pain severity, whereas other regions exhibited a mixture of positive and negative associations.

#### 3.2.2. Joint expression of pain severity and anxiety symptoms is associated with low- and high-frequency neural synchrony

PLSR revealed significant relationships between pain-anxiety composite score and neural synchrony across various brain networks and lobes (**Figure 2, Row 2**), primarily in the theta and gamma ranges.

For theta, higher pain-anxiety composite scores were predominantly associated with reduced neural synchrony, particularly in connections with the somatomotor network. In contrast, positive associations were observed for the connections between the medial prefrontal cortex and the right putamen, as well as between the medial prefrontal cortex and the left amygdala.

The magnitude of synchrony between the left pallidum to right putamen was also negatively associated with the pain-anxiety composite in the beta frequency band, representing the only connection to show a significant effect at this frequency.

For low-gamma1, many connections, especially those involving somatomotor regions, were negatively associated with pain-anxiety composite, whereas a smaller number of connections involving the ventral attention and subcortical networks showed positive associations. In the low-gamma2 frequency band, the connection between the mid-cingulate cortex and the right caudate was also negatively associated with pain-anxiety composite intensity. In high-gamma band, several functional connections across multiple networks exhibited positive and negative associations with pain-anxiety composite.

#### 3.2.3. Joint expression of pain severity and depressive symptoms is associated with wide-frequency neural synchrony across a limited set of connections

PLSR revealed significant associations between the pain-depression composite score and neural synchrony across most frequency bands, except for alpha and delta (**Figure 2**, **Row 3**).

In the theta band, higher pain-depression composite scores were inversely associated with synchrony between the right secondary motor cortex and the right secondary somatosensory cortex.

In the beta band, synchrony between the right putamen and left supplementary motor area was positively associated with the pain-depression composite, whereas connections between the right putamen and left pallidum, as well as between the right dorsolateral prefrontal cortex and left posterior insula were negatively associated.

In the low-gamma1 band, several within- and between-network connections involving somatomotor and subcortical regions were negatively associated with the pain-depression composite, in contrast to the positive association of this composite with synchrony between the right pallidum and left putamen. The connection between the mid-cingulate cortex and the right caudate was also negatively associated with pain-depression composite in the low-gamma2 frequency band. In the high-gamma, both positive and negative associations were observed across multiple regions across several brain networks.

#### 3.2.4 Joint expression of pain severity and PTSD-related symptoms is associated with neural synchrony in mid- to high-frequency bands

PLSR revealed significant associations between the pain-PTSD composite score and neural synchrony across the beta, low-gamma, and high-gamma frequency bands (**Figure 2**, **Row 4**).

In the beta band, synchrony between the left putamen and right dorsolateral prefrontal cortex was positively associated with the pain-PTSD composite. In contrast, connections from the left posterior insula to the right dorsolateral prefrontal cortex, from the right putamen to the left pallidum, and from the subgenual anterior cingulate cortex to the left occipital cortex were inversely associated with pain-PTSD composite intensity.

In the low-gamma1 band, several connections, especially those involving somatomotor, were negatively associated with the pain-PTSD composite. Conversely, a limited numbers of connections involving subcortical regions and other networks, as well as between the right secondary somatosensory cortex and the right temporoparietal junction, were positively associated with the pain-PTSD composite. In the high-gamma band, synchrony between many brain regions exhibited both positive and negative associations with pain-PTSD composite.

## 4. Discussion

### 4.1. Summary

In this study, we examined the relationship between frequency-specific neural synchrony and pain severity, as well as neural synchrony associated with the co-occurring expression of pain and psychological symptoms in the pain connectome **(Figure 2)**. Using Partial Least Squares Regression (PLSR) applied to MEG-based synchrony between selected brain regions in a military cohort with heterogenous pain severity and psychological symptom profiles, we addressed this transdiagnostic aim.

Using a transdiagnostic approach, we focused on a sample recruited from active military members and Veterans, a cohort characterized by heterogeneous chronic pain and mental health symptom profiles

We uncovered significant associations between pain severity and neural synchrony across multiple frequency bands, with the strongest effects observed in the beta and high-gamma bands across several brain networks, and more limited but significant associations in the theta and low-gamma bands.

Our results also revealed distinct patterns of neural synchrony associated with the joint expression of pain severity and psychological symptoms across frequency bands and brain networks. Co-occurring pain and anxiety symptoms were primarily associated with alterations in theta- and gamma-band synchrony, whereas co-occurring pain with depressive and PTSD-related symptoms was predominantly associated with gamma-band synchrony, with a smaller number of associations observed in the beta band.

Together, these findings demonstrate that pain severity and co-occurring psychological symptom burden are reflected in partially overlapping but frequency-specific neural synchrony patterns. This transdiagnostic, dimensional framework highlights how distributed brain network dynamics relate to shared symptom variability.

### 4.2. Comparisons with prior studies

Comparing our findings with electrophysiological studies on pain-related neural dynamics reveals both consistencies and contradictions across frequency bands and brain networks **(Figure 2, Row 1)**. Dinh et al. reported increased theta connectivity in frontal regions in individuals with elevated pain symptoms [51], aligning with our findings. Similarly, Kim et al. identified theta-band disruptions between the salience network and nociceptive pathways [29], supporting the notion that pain severity is associated with altered large-scale neural communication and impaired top-down modulation.

Kisler et al. reported increased alpha power in chronic pain patients [52], whereas we found no significant alpha synchrony effects. Similarly, Witjes et al. observed higher slow-to-fast alpha power ratios in multiple brain regions [53], absent in our results, possibly due to methodological differences between spectral power and phase-based synchrony metrics. Kim et al. also reported increased alpha coherence in the salience network [29], which we did not observe, suggesting variability based on methodology, population, or pain disorder type.

Beta-band synchrony exhibited more consistent overlap with prior work, reflecting its complex role in pain perception, cognitive control, and sensorimotor integration. Our findings showed significant beta-band associations involving salience, sensorimotor, and default mode networks, aligning with prior MEG studies. Jin et al. reported that conditioned pain modulation reduces sensorimotor beta activity [54], supporting its role in pain relief. In contrast, Diers et al. found beta activity increased in the insula and sensory cortex when attention was directed toward pain [55], highlighting its role in cognitive engagement and top-down modulation. Similarly, Ueno et al. reported altered beta connectivity in the sensorimotor and salience networks [30], reinforcing beta synchrony’s role in pain processing and cognitive-affective regulation.

Jin et al. demonstrated that conditioned pain modulation reduces sensorimotor beta activity [54], supporting the role of beta oscillations in pain regulation. In contrast, Diers et al. reported increased beta activity in insular and sensory cortices when attention was directed toward pain [55], highlighting beta-band involvement in cognitive engagement and top-down modulation. Ueno et al. similarly reported altered beta connectivity in sensorimotor and salience networks [30], reinforcing beta synchrony as a key component of pain-related neural dynamics.

Gamma-band synchrony showed the most robust and widespread associations with pain severity in our study, particularly in the high-gamma range. These findings are consistent with prior reports linking gamma oscillations to pain processing and cortical excitation, including increased frontal gamma connectivity in individuals with elevated pain symptoms [51]. Fauchon et al. suggests gamma oscillations may be more complex than assumed, with distinct subcomponents playing different roles in pain processing [28].

When comparing findings related to co-occurring pain and anxiety symptoms, our results align with prior work implicating theta-band synchrony in both pain and anxiety-related processes (**Figure 2**, **Row 2**). Increased theta connectivity has been reported in pain contexts [51], while hippocampal theta activity has been linked to anxiety [56]. In the beta band, our specific findings on the left pallidum-right putamen could not fully address reports of widespread beta alterations during threat [55] or increased beta activity in pain-related attention [57]. For gamma, our findings are consistent with evidence linking gamma-band activity to pain processing [28,51], and they are also compatible with prior work suggesting that gamma oscillations can vary with anxiety-related states and symptoms [58,59].

Our findings on association between neural synchrony and joint pain and depressive symptom burden show consistencies with prior research (**Figure 2**, **Row 3**). In the beta band, positive and negative associations in subcortical and prefrontal-insular connections align with altered beta connectivity in pain processing [30] and reduced beta functional connectivity in major depressive disorder [60], supporting its role in cognitive-affective regulation. In the gamma band, our results align with pain-related gamma connectivity [51], while depression-related hyperconnectivity in low-gamma between visual and limbic regions [61] further supports its role in affective processing.

Comparing our results with MEG studies on pain and PTSD-related symptoms provides further insights. Both symptom domains have been associated with altered beta- and gamma-band synchrony, with PTSD-related symptom severity linked to changes in neural connectivity [35,44,61,62] and pain severity showing similar frequency-specific alterations [51]. This aligns with our observed beta and gamma synchrony associations with joint pain-PTSD symptom burden (**Figure 2**, **Row 4**). In particular, higher-frequency oscillatory dynamics have been associated with PTSD symptom severity and network-level connectivity changes [35,61], mirroring our findings. Although theta-band alterations have been reported in both pain and PTSD contexts [63,29], we did not observe theta-band associations for joint pain-PTSD symptoms, suggesting frequency-specific dissociations across symptom domains.

### 4.3. Limitations & Future Direction

This study has several considerations that are important for interpreting the findings. Given the observational and cross-sectional nature of the design, the reported associations between frequency-specific neural synchrony, pain severity, and co-occurring psychological symptoms should be interpreted as correlational rather than causal. Our focus on dimensional symptom measures allowed us to characterize shared neural patterns across heterogeneous presentations, but the present findings do not establish temporal ordering or directionality of these brain-symptom relationships.

Future studies employing longitudinal and interventional designs will be essential to clarify how neural synchrony patterns evolve over time and whether they change in response to targeted treatments. Such approaches may help distinguish state-dependent from more stable neural features associated with pain and psychological symptom burden. Interventional studies, including pharmacological, behavioral, or neuromodulatory approaches, could further clarify whether modifying specific frequency-specific synchrony patterns leads to changes in pain or psychological symptoms.

Although the present study adopted a transdiagnostic, dimensional framework, future research could extend this work by examining clinically characterized patient populations with formally diagnosed comorbid pain and psychiatric conditions. Studying such cohorts may facilitate clearer clinical interpretation and help bridge the gap between dimensional neural markers and diagnosis-informed clinical applications.

Finally, future work integrating multimodal data, including neuroimaging, behavioral, and physiological measures, may provide a more comprehensive understanding of the mechanisms linking neural synchrony to pain and co-occurring psychological symptoms. Multimodal analytic approaches may help capture complementary aspects of neural and systemic processes that are not fully accessible through MEG alone.

### 4.4. Conclusion

The findings of this study provide insight into transdiagnostic neural synchrony patterns associated with pain severity and co-occurring psychological symptom burden, including anxiety-, depression-, and PTSD-related symptoms. By employing multivariate PLSR applied to MEG-based frequency-specific neural synchrony, we identified distributed patterns of neural coupling within the pain connectome that were associated with pain severity and its co-expression with psychological symptoms.

Our results demonstrate that pain severity and co-occurring psychological symptoms are reflected in partially overlapping but frequency-specific neural synchrony patterns across large-scale brain networks. Together, these findings identify shared, frequency-specific neural synchrony patterns underlying pain severity and psychological symptom burden, open new avenues for understanding pain-mental health comorbidity, and inform future multidimensional and transdiagnostic research and treatment efforts.

## Data Availability

The data supporting this study are not publicly available due to defence considerations and restrictions. However, to enhance transparency and reproducibility, we have shared scripts implementing the PLSR analyses and exploratory machine learning models at https://github.com/majidsaberi/PLSmodeling/

## Declaration of Generative AI Use

During the preparation of this work, the authors used AI-assisted tools to improve language and readability. The authors critically reviewed and edited all content and take full responsibility for the final manuscript.

