## Supplementary Material for "Neural synchrony reflects pain and co-occurring psychological symptoms: a transdiagnostic magnetoencephalography study using multivariate modeling"

| ROI Label | ROI Name | Network | Coordinate | | |
|---|---|---|---|---|---|
| | | | X | Y | Z |
| THAL.L | Left Thalamus | Subcortical | -12 | -18 | 8 |
| THAL.R | Right Thalamus | Subcortical | 12 | -18 | 8 |
| S1.L | Left Primary Somatosensory Cortex | Sensorimotor | -34 | -30 | 54 |
| S1.R | Right Primary Somatosensory Cortex | Sensorimotor | 34 | -28 | 54 |
| S2.L | Left Secondary Somatosensory Cortex | Sensorimotor | -60 | -30 | 20 |
| S2.R | Right Secondary Somatosensory Cortex | Sensorimotor | 60 | -22 | 18 |
| pINS.L | Left Posterior Insula | Sensorimotor | -34 | -20 | 18 |
| pINS.R | Right Posterior Insula | Sensorimotor | 34 | -20 | 18 |
| TPJ.L | Left Temporoparietal Junction | Ventral Attention | -50 | -42 | 28 |
| TPJ.R | Right Temporoparietal Junction | Ventral Attention | 50 | -32 | 28 |
| aINS.L | Left Anterior Insula | Salience | -34 | 18 | 4 |
| aINS.R | Right Anterior Insula | Salience | 34 | 18 | 4 |
| MCC.L | Left Mid Cingulate Cortex | Salience | 2 | 12 | 34 |
| dlPFC.L | Left Dorsolateral Prefrontal Cortex | Central Executive | -38 | 40 | 28 |
| dlPFC.R | Right Dorsolateral Prefrontal Cortex | Central Executive | 34 | 46 | 22 |
| PCC | Posterior Cingulate Cortex | Default Mode | -2 | -46 | 28 |
| mPFC | Medial Prefrontal Cortex | Default Mode | -2 | 50 | 2 |
| sgACC | Subgenual Anterior Cingulate Cortex | Limbic | 4 | 26 | -8 |
| AMYG.L | Left Amygdala | Limbic | -24 | -1 | -17 |
| AMYG.R | Right Amygdala | Limbic | 26 | 1 | -18 |
| M1.L | Left Primary Motor Cortex | Sensorimotor | -40 | -6 | 51 |
| M2.R | Right Secondary Motor Cortex | Sensorimotor | 40 | -8 | 52 |
| SMA.L | Left Supplementary Motor Area | Sensorimotor | -6 | 5 | 61 |
| SMA.R | Right Supplementary Motor Area | Sensorimotor | 8 | 0 | 62 |
| CAUD.L | Left Caudate | Subcortical | -12 | 11 | 9 |
| CAUD.R | Right Caudate | Subcortical | 14 | 12 | 9 |
| PUT.L | Left Putamen | Subcortical | -25 | 4 | 2 |
| PUT.R | Right Putamen | Subcortical | 27 | 5 | 2 |
| PALL.L | Left Pallidum | Subcortical | -19 | 0 | 0 |
| PALL.R | Right Pallidum | Subcortical | 20 | 0 | 0 |
| PreCUN | Precuneus | Default Mode | 2 | -61 | 48 |
| dmPFC | Dorsomedial Prefrontal Cortex | Default Mode | -13 | 52 | 23 |
| OCC.L | Left Occipital Cortex | Visual | -14 | -94 | 24 |
| OCC.R | Right Occipital Cortex | Visual | 10 | -94 | 24 |
| MTL.L | Left Medial Temporal Lobe | Default Mode | -62 | -22 | 8 |
| MTL.R | Right Medial Temporal Lobe | Default Mode | 58 | -22 | 8 |

**Supplementary Table 1.** Regions of interest for the dynamic pain connectome, their coordinates, and the brain networks to which they belong.
